# Antibiotic Timing and Survival After Immune Checkpoint Inhibitor Initiation in Patients With Cancer

**DOI:** 10.64898/2026.05.27.26354193

**Authors:** Kevin Zhang, Daniel John, Wei Tse Li, Michael Hogarth, Rana R. McKay, Weg M. Ongkeko

## Abstract

**Importance:** While gut dysbiosis is known to impair response to immune checkpoint inhibitors (ICIs), the relative clinical impact of antibiotic timing (pre-vs. post-ICI initiation) remains unclear.

**Objective:** To evaluate whether antibiotic timing differentially influences overall survival (OS) in a large, multi-institutional pan-cancer cohort.

**Design, Setting, and Participants:** This retrospective cohort study utilized deidentified electronic health record data from six academic medical centers within the University of California Health system. We included 21,108 adults with any malignancy who received PD-1, PD-L1, or CTLA-4 inhibitors between January 2014 and December 2024.

**Exposures:** Antibiotic exposure windows were categorized as pre-only (−60 to −1 days), post-only (+1 to +60 days), both windows, or none.

**Main Outcomes and Measures:** The primary outcome was overall survival (OS) calculated from the first ICI dose. Multivariable Cox proportional hazards models adjusted for demographics, tumor type, line of therapy, and baseline health indicators (albumin, NLR, and recent hospitalization).

**Results:** Among 21,108 patients, 17.3% had pre-only exposure, 13.3% had post-only exposure, and 60.6% had no exposure. In the multivariable model, post-only exposure (HR, 1.27; 95% CI, 1.20–1.35) and combined pre- and post-exposure (HR, 1.31; 95% CI, 1.23–1.40) were significantly associated with higher mortality. Pre-only exposure was not significantly associated with OS (HR, 1.04; 95% CI, 0.99–1.10). Subgroup analyses by tumor type showed consistent trends across major malignancies, including head and neck (Post HR, 1.46) and renal cell carcinoma (Post HR, 1.26).

**Conclusions and Relevance:** In contrast to some smaller studies, this large-scale analysis indicates that antibiotic exposure *after* ICI initiation carries a greater risk than exposure *prior* to treatment. These findings highlight the need for rigorous antibiotic stewardship strategies specifically during the early phases of immunotherapy treatment.

**Key Points:** 

**Question:** Does the timing of antibiotic administration relative to immune checkpoint inhibitor (ICI) initiation differentially affect overall survival across a pan-cancer cohort?

**Findings:** In this retrospective cohort study of 21,108 patients, antibiotic exposure within 60 days *after* starting ICIs was associated with a 27% increased risk of death (HR, 1.27; 95% CI, 1.20-1.35). Conversely, exposure occurring *only* before ICI initiation showed no significant long-term survival association in adjusted models (HR, 1.04; 95% CI, 0.99-1.10).

**Meaning:** Clinical focus should shift toward antibiotic stewardship during the active phase of immunotherapy, as post-initiation exposure appears to be a more critical driver of poor outcomes than pre-treatment exposure.

## Introduction

Antibiotic exposure and immune checkpoint inhibitor (ICI) therapy are both common in cancer care, with various outcomes observed. ICIs are standard across cancers, with efficacy shaped by host immune and microbial factors.^1,9^ Antibiotics disrupt the gut microbiome, and a systematic review confirmed that antibiotic use is associated with worse survival in ICI-treated patients, although timing windows varied greatly.^2,8,10^

Several studies suggest detrimental effects of pre-ICI antibiotics,^3^,^4^ while others examined post-ICI use.^5^ A large cohort found no unique association between pre-ICI antibiotics and survival after accounting for confounding variables.^6^ No prior pan-cancer study has directly assessed the association of mutually exclusive pre-only and post-only exposure of antibiotics with ICI outcomes. We therefore evaluated whether antibiotic timing is associated with differential survival in a large, multi-institutional cohort of 11 primary cancer types.

## Methods

We conducted a retrospective cohort study using IRB-approved, deidentified electronic health record data from the University of California Health Data Warehouse, which integrates records from six academic medical centers across California. Adults with any malignancy who received at least one dose of PD-1, PD-L1, or CTLA-4 inhibitor between January 2014 and December 2024 were included. The index date was the first ICI administration. We included only systemic oral or intravenous antibiotics with known microbiome impact (penicillins, cephalosporins, carbapenems, fluoroquinolones, macrolides, tetracyclines, sulfonamides, metronidazole, clindamycin, vancomycin, linezolid). To avoid counting prophylactic or incidental use, we required ≥2 distinct days of administration within the exposure window. Exposure was categorized as pre-only (−60 to −1 days before ICI start), post-only (+1 to +60 days after ICI start), or none; patients with antibiotics in both windows were retained as a separate category in sensitivity analyses. Covariates included demographics, tumor type, line of therapy, treatment site, albumin, neutrophil-to-lymphocyte ratio, corticosteroid use, and recent hospitalization. The primary outcome was overall survival. Kaplan–Meier curves and multivariate Cox models with Schoenfeld residuals were used. Analyses were conducted in Python 3.10 (lifelines package).

## Results

Among 21,108 patients, 12,785 (60.6%) had no antibiotic exposure, 3,657 (17.3%) had pre-only, and 2,805 (13.3%) had post-only. Baseline characteristics revealed that the pre-only group had the highest clinical severity markers, including a 42.4% rate of prior hospitalization compared to 27.0% in the post-only group and 21.2% in the no-antibiotic group (Table 1).

**Table 1.** Baseline Demographic and Clinical Characteristics of Patients Stratified by Antibiotic Exposure Timing.

| Characteristic | None (n=16,504) | Pre Only (n=3,716) | Post Only (n=3,183) | Both (n=1,810) |
| --- | --- | --- | --- | --- |
| **Gender** |  |  |  |  |
| Female | 7468 (45.2%) | 1643 (44.2%) | 1489 (46.8%) | 776 (42.9%) |
| Male | 9034 (54.7%) | 2071 (55.7%) | 1694 (53.2%) | 1034 (57.1%) |
| Unknown | 2 (0.0%) | 2 (0.1%) | 0 (0.0%) | 0 (0.0%) |
| **Race** |  |  |  |  |
| White | 10252 (62.1%) | 2275 (61.2%) | 1968 (61.8%) | 1085 (59.9%) |
| Asian | 2139 (13.0%) | 488 (13.1%) | 393 (12.3%) | 266 (14.7%) |
| Other Race | 1997 (12.1%) | 464 (12.5%) | 413 (13.0%) | 239 (13.2%) |
| Black or African American | 551 (3.3%) | 150 (4.0%) | 106 (3.3%) | 72 (4.0%) |
| Multirace | 506 (3.1%) | 107 (2.9%) | 110 (3.5%) | 56 (3.1%) |
| Native Hawaiian/Pacific Islander | 66 (0.4%) | 24 (0.6%) | 19 (0.6%) | 8 (0.4%) |
| American Indian/Alaska Native | 50 (0.3%) | 8 (0.2%) | 12 (0.4%) | 2 (0.1%) |
| Unknown/Other Categories | 943 (5.7%) | 200 (5.4%) | 162 (5.1%) | 82 (4.5%) |
| **Cancer Category** |  |  |  |  |
| Lung Cancer | 2486 (15.1%) | 677 (18.2%) | 512 (16.1%) | 301 (16.6%) |
| Other Solid Tumors<sup>a</sup> | 3397 (20.6%) | 808 (21.7%) | 641 (20.1%) | 425 (23.5%) |
| Breast | 1350 (8.2%) | 206 (5.5%) | 245 (7.7%) | 65 (3.6%) |
| Melanoma / Skin | 1228 (7.4%) | 242 (6.5%) | 197 (6.2%) | 93 (5.1%) |
| Renal Cell Carcinoma | 1163 (7.0%) | 197 (5.3%) | 202 (6.3%) | 97 (5.4%) |
| Head and Neck | 857 (5.2%) | 238 (6.4%) | 205 (6.4%) | 182 (10.1%) |
| Gastric / Esophageal | 892 (5.4%) | 169 (4.5%) | 141 (4.4%) | 74 (4.1%) |
| Gynecologic | 1175 (7.1%) | 234 (6.3%) | 236 (7.4%) | 103 (5.7%) |
| Liver / HCC | 838 (5.1%) | 192 (5.2%) | 171 (5.4%) | 91 (5.0%) |
| Urothelial / Bladder | 764 (4.6%) | 264 (7.1%) | 219 (6.9%) | 153 (8.5%) |
| Colorectal | 571 (3.5%) | 151 (4.1%) | 123 (3.9%) | 64 (3.5%) |
| Unknown | 1783 (10.8%) | 338 (9.1%) | 291 (9.1%) | 162 (9.0%) |
| **Clinical Factors** |  |  |  |  |
| Previous Systemic Therapy (Yes) | 3298 (20.0%) | 888 (23.9%) | 659 (20.7%) | 482 (26.6%) |
| Prior Hospitalization (Yes) | 3505 (21.2%) | 1576 (42.4%) | 859 (27.0%) | 844 (46.6%) |
| Corticosteroid Use (Yes) | 4923 (29.8%) | 1286 (34.6%) | 1198 (37.6%) | 710 (39.2%) |
| **Laboratory Values** |  |  |  |  |
| Age, Median [IQR] | 67.0 [57.0-75.0] | 67.0 [57.0-75.0] | 67.0 [57.0-75.0] | 66.0 [56.0-75.0] |
| Albumin (g/dL), Median [IQR] | 3.9 [3.5-4.2] | 3.7 [3.3-4.1] | 3.8 [3.3-4.1] | 3.6 [3.1-4.0] |
| Neutrophils, Median [IQR] | 4.4 [3.2-6.2] | 5.0 [3.6-7.4] | 4.8 [3.4-6.9] | 5.3 [3.7-8.0] |
| Lymphocytes, Median [IQR] | 1.2 [0.8-1.7] | 1.2 [0.8-1.6] | 1.1 [0.7-1.6] | 1.1 [0.7-1.6] |
| **Primary Study Site** |  |  |  |  |
| UCLA | 5360 (32.5%) | 1273 (34.3%) | 1099 (34.5%) | 635 (35.1%) |
| UCSF | 3705 (22.4%) | 765 (20.6%) | 691 (21.7%) | 437 (24.1%) |
| UCSD | 3486 (21.1%) | 808 (21.7%) | 668 (21.0%) | 349 (19.3%) |
| UCI | 2325 (14.1%) | 556 (15.0%) | 444 (13.9%) | 236 (13.0%) |
| UCD | 1623 (9.8%) | 313 (8.4%) | 279 (8.8%) | 153 (8.5%) |
| UCR | 5 (0.0%) | 1 (0.0%) | 2 (0.1%) | 0 (0.0%) |

<sup>a</sup> Other Solid Tumors includes various primary neoplasms including, but not limited to, Sarcoma, Pancreatic, Ovarian, and Prostate cancer] and other rare tumor types

In the first 100 days after ICI initiation, the pre-only group experienced the steepest early decline in survival (Figure 1A). However, over 5 years of follow-up, the post-only group survival curve diverged and showed the poorest long-term outcomes (Figure 1B) Median overall survival was longest in patients with no antibiotic exposure (428 days), followed by those in the pre-only (358 days) and post-only (338 days) groups, while patients with antibiotic exposure in both windows had the shortest median survival (281 days).

**Fig 1.**
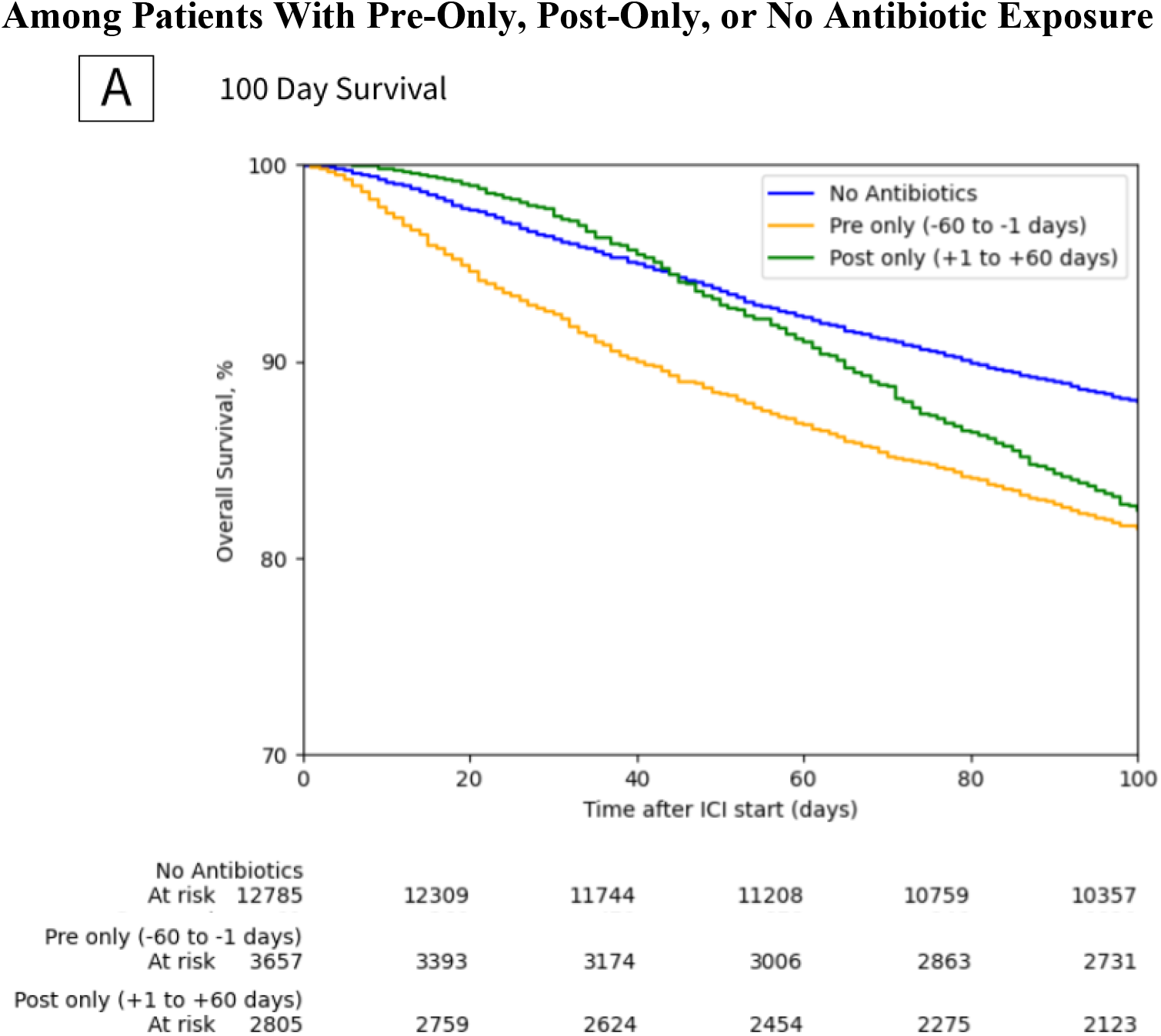
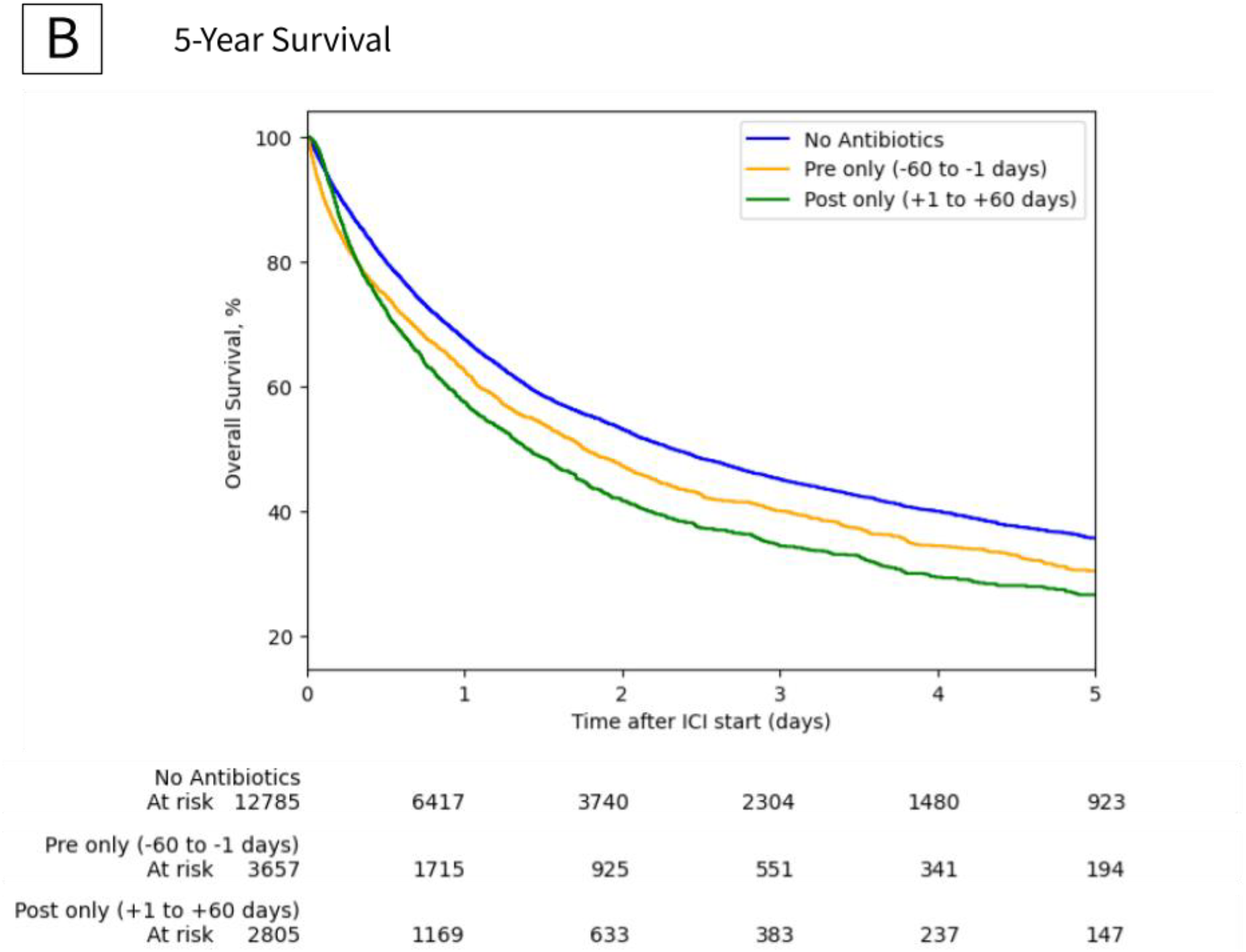
Overall Survival Rates Within 100 Days and Up to 5 Years After Immune Checkpoint Inhibitor Initiation. The curves show overall survival among patients who received pre-only or post-only antibiotics compared with those without antibiotic exposure. Early mortality was higher in the pre-only group, whereas over longer follow-up the post-only group showed poorer survival (log-rank [Mantel-Cox] test, P < .001).

In multivariable analysis, post-only exposure (HR, 1.27; 95% CI, 1.20–1.35) and exposure in both windows (HR, 1.31; 95% CI, 1.23–1.40) were significantly associated with mortality. Pre-only exposure was not significant (HR, 1.04; 95% CI, 0.99–1.10). In subgroup analyses stratified by tumor type, every individual cancer category was limited by small antibiotic exposure groups, often consisting of fewer than 300 patients per window. Despite these sample size constraints, a significant survival disadvantage for post-only antibiotic exposure was identified in head and neck cancer (HR, 1.46; 95% CI, 1.20–1.77), renal cell carcinoma (HR, 1.26; 95% CI, 1.03–1.54), and breast cancer (HR, 1.24; 95% CI, 1.01–1.54). However, many other common malignancies, including lung cancer (HR, 1.09; 95% CI, 0.97–1.23) and melanoma (HR, 1.07; 95% CI, 0.85– 1.34), did not reach statistical significance, exhibiting modest effect sizes and wide confidence intervals that likely reflect the limited power within these specific exposure windows (Figure 2).

**Fig 2.**
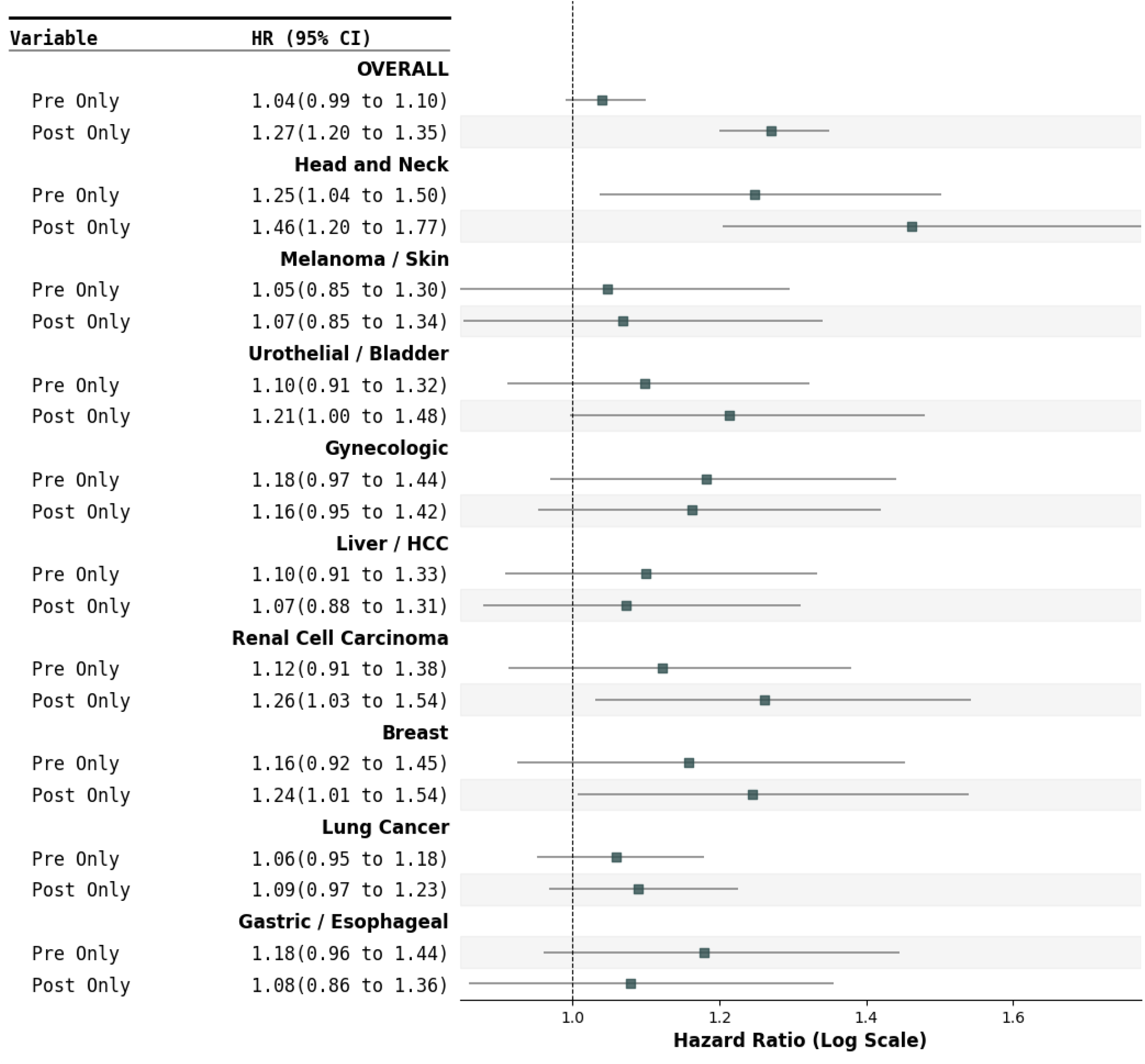
Multivariable-Adjusted Hazard Ratios for Mortality Among Patients With Pre-Only or Post-Only Antibiotic Exposure Stratified by Cancer Type. The forest plot shows the adjusted risk of mortality among patients who received pre-only or post-only antibiotics compared with those without antibiotic exposure. Across the overall cohort and major cancer subtypes, post-only exposure (+1 to +60 days) was associated with significantly higher hazard ratios for mortality compared with pre-only exposure (−60 to −1 days), which did not show a significant long-term association with survival in the multivariable model.

## Discussion

In this multi-institutional study of 21,108 patients, we demonstrate that the chronological relationship between antibiotic (ATB) exposure and the initiation of immune checkpoint inhibitors (ICIs) is a significant predictor of overall survival. While existing literature has primarily investigated the role of prior antibiotic exposure, our data indicate that ATB therapy administered within the first 60 days following ICI commencement (post-only) carries a significantly stronger association with increased mortality. Specifically, post-only exposure was independently associated with a 27% increase in mortality risk (HR, 1.27; 95% CI, 1.20–1.35), whereas pre-only exposure did not reach statistical significance in our multivariable model (HR, 1.04; 95% CI, 0.99–1.10). While our findings are correlational and do not definitively establish causality, the distinct survival disadvantage observed specifically in the post-ICI window suggests a time-dependent biological interaction between the gut microbiome and early T-cell activation.

Our subgroup analysis suggests that the impact of antibiotics is highly dependent on tumor biology. Notably, our findings for lung cancer (HR, 1.09; 95% CI, 0.97–1.23) deviate from previous literature that characterized this malignancy as particularly susceptible to antibiotic-induced ICI failure^3,4,9^. Despite representing our largest cohort (n=3,976), the survival disadvantage in this group was modest and non-significant, potentially indicating a higher degree of clinical resilience to microbial disruption in real-world lung cancer populations than previously recognized. Furthermore, our null findings for melanoma (HR, 1.07) are consistent with the results reported by Poizeau et al^11^.

Conversely, significant survival deficits for post-only exposure were observed in head and neck cancer (HR, 1.46), renal cell carcinoma (HR, 1.26), and breast cancer (HR, 1.24). The significant association found in our breast cancer cohort notably aligns with findings by Kulkarni et al^5^. We hypothesize that these malignancies may be more “microbiome-sensitive” due to their typically high levels of immune cell infiltration. Exposure to antibiotics during the 60-day post-initiation window may be particularly detrimental in these “hot” tumor microenvironments because it coincides with the critical expansion and priming of tumor-reactive T-cells. This disruption may deprive the immune system of essential microbial-derived signals exactly when pharmacological activation is intended to occur.

The “prior hospitalization paradox” observed in our cohort further suggests that these outcomes are not merely a reflection of baseline clinical frailty. Patients in the pre-only group exhibited higher baseline severity, including a prior hospitalization rate double that of the post-only group (42.4% vs. 27.0%). Despite appearing clinically “healthier” at treatment onset, the post-only group demonstrated inferior survival outcomes (338 days vs. 358 days median survival). This divergence suggests that while the immune-microbiome axis may possess resilience to disruptions prior to ICI therapy, the early post-initiation phase represents a “critical window” where microbial integrity is vital for therapeutic efficacy.

In conclusion, our results indicate that clinical antibiotic stewardship should be nuanced and potentially personalized by tumor type. While a negative survival trend exists across the broader cohort, the clinical significance of antibiotic exposure appears dictated by specific oncologic contexts and the precise timing of the dose.

### Limitations

This study is limited by its retrospective, correlational design, which precludes establishing direct causality. We lacked gut microbiota data and analysis (e.g., *Akkermansia* or *Bifidobacterium*), as well as granular EHR data on ECOG status and PD-L1 expression. Additionally, evidence of nonproportional hazards for certain covariates, such as age and albumin, suggest their survival influence may vary over time. Furthermore, individual subtypes were limited by small exposure groups (often n < 300), resulting in wider confidence intervals for less prevalent cancers. Despite these factors, the scale of this multi-institutional analysis of over 21,000 unselected patients ensures high external validity.

## Conclusion

This pan-cancer analysis addresses a critical gap in the literature by explicitly disentangling the survival impact of antibiotic exposure windows relative to ICI initiation. Our data demonstrate that the first 60 days of immunotherapy represent a period of heightened vulnerability, where antibiotic use is a significantly more potent driver of mortality than pre-treatment exposure. These findings suggest that clinical antibiotic stewardship should not be limited to baseline screening but must be prioritized as a continuous intervention during the active phase of treatment. Prospective trials are now needed to determine if preserving or restoring the gut microbiome specifically during this 60-day window can improve the durability of immunotherapy and overall patient survival.

## Data Availability

All data produced in the present study are available upon reasonable request to the authors.

